# Phenome-wide association analysis of substance use disorders in a deeply phenotyped sample

**DOI:** 10.1101/2022.02.09.22270737

**Authors:** Rachel L. Kember, Emily E. Hartwell, Heng Xu, James Rotenberg, Laura Almasy, Hang Zhou, Joel Gelernter, Henry R. Kranzler

## Abstract

**Background:** Substance use disorders (SUDs) are associated with a variety of co-occurring psychiatric disorders and other SUDs, which partly reflects genetic pleiotropy. Polygenic risk scores (PRS) and phenome-wide association studies (PheWAS) are useful in evaluating pleiotropic effects. The comparatively low prevalence of SUDs and lack of detailed information available in electronic health records limits their informativeness for such analyses.

**Methods:** We used the deeply-phenotyped Yale-Penn sample [(N=10,610; 46.3% African ancestry (AFR), 53.7% European ancestry (EUR)], recruited for genetic studies of substance dependence, to examine pleiotropy for 4 major substance-related traits: alcohol use disorder (AUD), opioid use disorder (OUD), smoking initiation (SMK), and lifetime cannabis use (CAN). The sample includes both affected and control subjects interviewed using the Semi-Structured Assessment for Drug Dependence and Alcoholism (SSADDA), a comprehensive psychiatric interview.

**Results:** In AFR individuals PRS for AUD, and in EUR individuals PRS for AUD, OUD, and SMK, were associated with their respective primary DSM diagnoses. These PRS were also associated with additional phenotypes involving the same substance. PheWAS analyses of PRS in EUR individuals identified associations across multiple phenotypic domains, including phenotypes not commonly assessed in PheWAS analyses, such as family environment and early childhood experiences.

**Conclusions:** Smaller, deeply-phenotyped samples can complement large biobank genetic studies with limited phenotyping by providing greater phenotypic granularity. These efforts allow associations to be identified between specific features of disorders and genetic liability for SUDs, which help to inform our understanding of the pleiotropic pathways underlying them.

## Introduction

Individuals with substance use disorders (SUDs) are at increased risk of comorbid psychiatric and medical disorders(1). Comorbidity of disease poses a challenge to both the diagnosis and treatment of SUDs, but the etiologic factors underlying comorbidity are not well understood. Large-scale genome-wide association studies (GWAS) have identified common risk markers for many SUDs(2–6) and established a pattern of genetic correlations among SUDs and between SUDs and other traits. This growing body of evidence suggests that there are common loci or biological pathways that contribute to the risk for multiple SUDs and psychiatric disorders. Identifying these pleiotropic loci and pathways could provide insight into the etiologies of co-occurring disorders and thereby advance efforts to categorize, prevent, and treat SUDs and co-occurring medical and psychiatric conditions.

The large samples required to identify variants of generally small effect often necessitate that the phenotypic information collected on participants be neither purpose-collected nor very detailed. This trade-off between sample size and depth of phenotyping limits clinically meaningful insights into disease biology(7). Furthermore, while the genome scan is hypothesis-free, the selection of a phenotype for GWAS – if not chosen of necessity because of limited phenotype data available – requires that assumptions be made regarding the most representative or informative traits. For this reason, phenome scans(8), also known as phenome-wide association studies (PheWAS)(9), are complementary to GWAS as they test the phenome in a similar hypothesis-free manner.

PheWAS have been most commonly implemented using data from electronic health records (EHR), where International Classification of Disease (ICD) codes can be converted to a simplified dataset that contains case/control status for over 1,800 diseases(9). A recent PheWAS of genetic liability for SUDs, represented by polygenic risk scores (PRS), identified cross-trait associations across multiple phenotypic domains in EHR data(10). However, PheWAS that use EHR data, as with GWAS that use EHR data, are limited by their reliance on ICD diagnoses and minimal phenotyping. PheWAS have also been performed on data extracted from epidemiological studies or clinical trials(11,12), an approach that allows testing of subthreshold (with respect to diagnosis) and non-diagnosis-based phenotypes.

The Yale-Penn sample, recruited for genetic studies of SUDs, was deeply phenotyped using the Semi-Structured Assessment for Drug Dependence and Alcoholism (SSADDA). This comprehensive psychiatric interview schedule assesses the physical, psychosocial, and psychiatric manifestations of SUDs and co-occurring psychiatric disorders(13,14). It includes >3500 items, and queries demographic information, lifetime diagnostic criteria for DSM-IV(15) and DSM-5(16) SUDs and DSM-IV(15) psychiatric disorders, psychosocial history, medical history, and a detailed history of substance use. The SSADDA yields reliable diagnoses and criterion counts for SUDs and psychiatric disorders(13,14).

The highly detailed information available on the Yale-Penn sample offers the opportunity for insights into the shared genetic etiology of a variety of substance use and psychiatric traits. The Yale-Penn dataset has been used to conduct genome-wide association studies(17–21), gene x environment studies(22,23), and phenotypic investigations(24,25). Although most of the literature on the genetics of SUDs derives from studies of individuals of European ancestry (EUR), the Yale-Penn dataset includes nearly equal numbers of EUR and African ancestry (AFR) individuals, so that analyses can be conducted in both ancestral groups.

Here we describe the selection of a subset of the data points collected with the SSADDA to create a PheWAS dataset based on the Yale-Penn sample. Using PRS for alcohol use disorder (AUD), opioid use disorder (OUD), smoking initiation (SMK), and lifetime cannabis use (CAN), we demonstrate the utility of this dataset for evaluating SUD pleiotropy. Further, we identify associations that contribute to our understanding of the shared genetic etiology and phenotypic comorbidity of these traits.

## Methods

### Yale-Penn dataset

Yale-Penn participants (N=16,715) were recruited at 5 sites in the United States for genetic studies of cocaine, opioid, and alcohol dependence. The study was approved by the institutional review board at each site and all participants gave written informed consent prior to data collection. Participants recruited as cases were identified through addiction treatment facilities, inpatient and outpatient psychiatric services, and advertisements in local media, and were screened for the presence of ≥ 1 of the 3 SUD diagnoses. Some cocaine-or opioid-dependent individuals were recruited as probands of small nuclear families and when available their affected and unaffected parents and siblings were also recruited. Unaffected controls were recruited from non-psychiatric medical settings and through advertisements.

### Semi-Structured Assessment for Drug Dependence and Alcoholism

The SSADDA is a comprehensive psychiatric interview that yields up to 3,727 data points (depending on skip patterns). It comprises 24 modules that assess the physical, psychological, social, and psychiatric manifestations of SUDs, psychiatric disorders, and environmental covariates considered likely to have an impact on SUDs. Administered as a computer-assisted interview, the SSADDA allows direct entry of subjects’ responses by the interviewer.

Demographic information elicited with the SSADDA includes sex, age, height, weight, education, employment, and relationship status. Environmental variables that were considered likely to have an impact on the risk of alcohol and drug dependence, including adverse childhood experiences, are also assessed. Medical history for common diseases is assessed by a yes/no response to “Have you been diagnosed with…” and followed by a series of medical disorders that the participant is asked to endorse.

The SSADDA’s semi-structured format, accompanied by rigorous training and quality control procedures(13), allow a carefully trained non-clinician interviewer to assess diagnostic criteria and disorders. It queries age of symptom onset, severity, duration and craving for the major drugs of abuse that yield DSM-IV diagnoses of nicotine dependence; dependence or abuse for other substances; mood disorders [major depressive disorder (MDD); bipolar disorder]; schizophrenia; mania; conduct disorder; antisocial personality disorder (ASPD); attention deficit hyperactivity disorder (ADHD); suicidality; anxiety disorders [panic disorder, agoraphobia, social anxiety disorder, obsessive-compulsive disorder, generalized anxiety disorder, posttraumatic stress disorder (PTSD)]; and gambling disorder. Recoding of criteria to accord with DSM-5 also makes it possible to generate DSM-5 SUD, but not psychiatric diagnoses.

### Variable selection and data cleaning

We used an interactive process among study clinicians (EEH, JR, HRK), a data scientist (HX), and a geneticist (RLK) to reduce the number of variables to 685 for use in PheWAS. Variables were retained if there was a consensus among the raters that they were informative for genetic studies and non-duplicative. Following variable selection, the data were cleaned to ensure consistency across categories. Full details of variable selection and data cleaning are available in Supplementary methods.

### Case and control definitions

Participants who met diagnostic criteria for a disorder were coded as “cases” and those who met no diagnostic criteria for the disorder were coded as “controls”. Subthreshold cases, for example those meeting at least one, but less than the required number of criteria for a diagnosis, were excluded from further analyses for that disorder. For symptom variables, participants who endorsed an individual symptom were considered cases and those who did not were controls. If an item was not answered, it was coded as “NA” and individuals were considered neither case nor control for that phenotype.

### Genotyping, imputation and Polygenic Risk Scores

Yale-Penn samples were genotyped in 3 batches using the Illumina HumanOmni1-Quad microarray, the Illumina HumanCoreExome array, or the Illumina Multi-Ethnic Global array. Genotyping quality control has been described in detail previously(19,20,26). Genotype data were imputed using the Michigan Imputation Server(27) with the 1000 Genomes Phase 3 reference panel(28). Further details are available in Supplementary methods.

Polygenic risk scores (PRS) were calculated for AUD(5), OUD(3), SMK(2), and CAN(4) using Polygenic Risk Scores–Continuous Shrinkage software(29) (PRS-CS) (Supplementary Table 1). We used the PRS-CS “auto” option to estimate the parameters of shrinkage priors and fixed the random seed to 1 for replicable results. When available, we used matched ancestry summary statistics (e.g., an AFR GWAS for AUD was used to calculate AUD PRS in AFR Yale-Penn individuals).

### Statistical Analysis

We conducted PheWAS of the 4 PRSs by fitting logistic regression models for binary traits and linear regression models for continuous traits. For both analytic models, we adjusted for sex, age, and the top 10 PCs within each genetic ancestry. Binary phenotypes with fewer than 100 cases or 100 controls and continuous phenotypes with fewer than 100 individuals within each ancestral group were excluded for that group. A Bonferroni correction was applied within each ancestral group to account for multiple testing (AFR p<8.7×10^−5^, EUR p<7.9×10^−5^).

## Results

### Sample

The PheWAS dataset comprises 685 variables in 25 phenotypic categories: 8 for substance use, 14 for psychiatric disorder, and 3 (demographics, environment, medical) for other features. Table 1 shows the demographic and clinical features of the analytic sample, and Supplementary Table 2 shows the case counts for all diagnoses. The sample (N=10,610) was majority male (overall: 55.6%; AFR: 54.9%; EUR: 56.2%) and included 4,918 AFR participants (996 or 20.3% with no SUD diagnosis) and 5,692 EUR participants (1,379 or 24.2% with no SUD diagnosis). The mean number of SUD diagnoses in the sample was 2.39 (SD=1.89) for DSM-IV and 2.28 (SD=1.82) for DSM-5. In this study, we focused on individuals with ≧1 SUD diagnoses for 4 substances [DSM-IV alcohol dependence (AD), opioid dependence (OD), tobacco dependence (TD), or cannabis dependence (CD), or DSM-5 alcohol use disorder (AUD), opioid use disorder (OUD), or cannabis use disorder (CUD)], comprising 3,813 AFR (38.7% female) and 4,287 EUR (37.2% female) individuals [average age = 40.1 (SD=10.9)]. There is a high degree of co-occurrence of SUD diagnoses in both population groups (Figure 1, Supplementary Tables 3-5).

**Table 1:** Demographic and Clinical Features

| Table 1: Demographic and Clinical Features |  |  |  |  |  |  |  |  |  |  |  |  |  |  |  |
| --- | --- | --- | --- | --- | --- | --- | --- | --- | --- | --- | --- | --- | --- | --- | --- |
| Ancestry |  | Diagnosis |  |  |  |  |  |  |  |  |  |  |  |  |  |
|  |  | DSM-IV |  |  |  |  |  |  |  | DSM-5 |  |  |  |  |  |
|  |  | Tobacco Dependence |  | Alcohol Dependence |  | Opioid Dependence |  | Cannabis Dependence |  | Alcohol Use Disorder |  | Opioid Use Disorder |  | Cannabis Use Disorder |  |
|  |  | N (% female) | Age (mean±SD) | N (% female) | Age (mean±SD) | N (% female) | Age (mean±SD) | N (% female) | Age (mean±SD) | N (% female) | Age (mean±SD) | N (% female) | Age (mean±SD) | N (% female) | Age (mean±SD) |
| African | Cases | 2794 (40.3) | 42.2 ± 9.2 | 2623 (35.6) | 42.0 ± 9.6 | 940 (35.2) | 44.5 ± 8.7 | 1406 (28.5) | 40.3 ± 9.8 | 3166 (36.2) | 41.7 ± 9.6 | 1017 (34.9) | 44.5 ± 8.7 | 2085 (30.8) | 40.7 ± 9.7 |
|  | Controls | 1522 (57.6) | 39.1 ± 11.6 | 1470 (64.2) | 40.7 ± 11.4 | 3844 (47.9) | 40.5 ± 10.4 | 2532 (56.0) | 41.7 ± 10.6 | 1413 (65.2) | 40.6 ± 11.5 | 3834 (48.1) | 40.5 ± 10.4 | 2376 (58.2) | 41.7 ± 10.7 |
| European | Cases | 3257 (37.7) | 38.2 ± 11.7 | 2935 (34.1) | 39.1 ± 11.6 | 2278 (34.7) | 36.5 ± 10.4 | 1631 (28.8) | 35.1 ± 10.7 | 3600 (34.5) | 38.7 ± 11.7 | 2404 (34.2) | 36.5 ± 10.5 | 2417 (28.7) | 36.2 ± 11.1 |
|  | Controls | 1963 (54.7) | 42.1 ± 14.7 | 1810 (61.2) | 42.0 ± 15.0 | 3240 (51.0) | 42.1 ± 14.2 | 3078 (55.1) | 42.5 ± 13.8 | 1639 (62.4) | 42.6 ± 15.1 | 3199 (51.5) | 42.1 ± 14.2 | 2842 (57.1) | 42.7 ± 14.0 |

**Figure 1.**
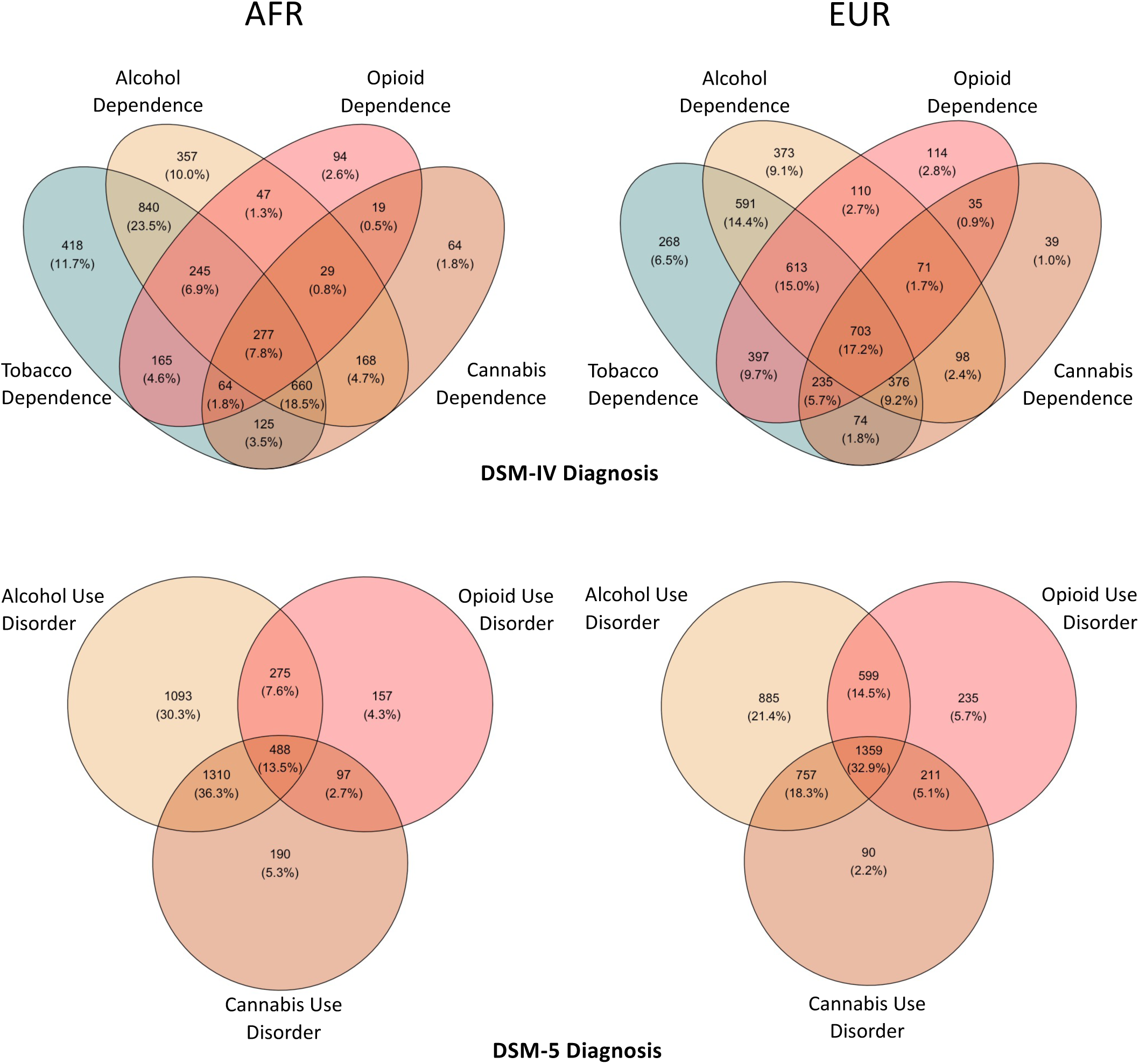
Comorbidity among SUDs in the Yale-Penn dataset. Venn diagrams show the comorbidity among DSM-IV substance dependence diagnoses (top) and DSM-5 use disorder diagnoses (bottom) in AFR individuals (left) and EUR individuals (right). Number and percentage of individuals with overlapping disorders is labelled in each section.

### Primary Associations of SUD PRS

In AFR individuals, PRS for AUD (PRS_AUD_) was more significantly associated with the DSM-5 AUD criterion count (β=0.30, p=5.4×10^−7^) than either the DSM-IV AD (OR=1.20, p=7.0×10^−6^) or the DSM-5 AUD diagnosis (OR=1.21, p=1.8×10^−6^; Figure 2A, Supplementary Table 6). PRS for OUD (PRS_OUD_) and smoking initiation (PRS_SMK_) were not significantly associated with either of their respective diagnoses or any other phenotypes (Figure 2A, Supplementary Tables 7 and 8). We were unable to generate PRS for cannabis lifetime use (PRS_CAN_) in AFR individuals as the discovery data were limited to EUR ancestry.

**Figure 2.**
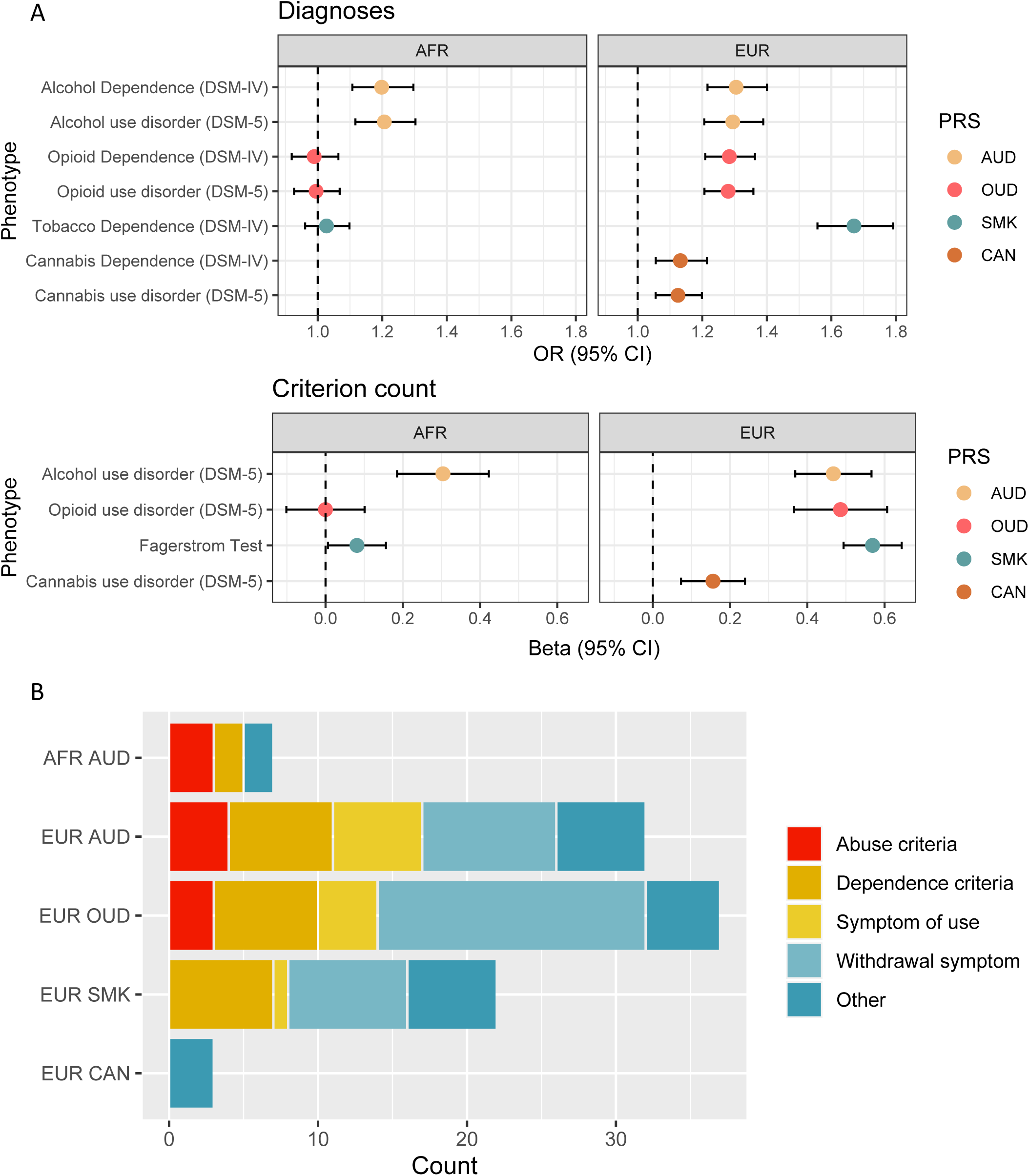
SUD PRS primary associations. A. Forest plot showing odds ratios and 95% confidence intervals for associations between each PRS and the primary DSM-IV and DSM-5 diagnosis in AFR individuals (left) and EUR individuals (right). B. Stacked bar plot showing the number and type of phenotypes involving the same substance associated with each PRS.

In EUR individuals, PRS_AUD_ and PRS_OUD_ were significantly associated with their respective DSM-IV and DSM-5 diagnoses and DSM-5 criterion counts (Figure 2A, Supplementary Tables 9 and 10). Similarly, PRS_SMK_ was significantly associated with the DSM-IV diagnosis of TD and FTND score (Figure 2A, Supplementary Table 11). The association of PRS_AUD_ with the DSM-5 criterion count was orders of magnitude more significant (β=0.47, p=2.3×10^−20^) than with either the DSM-IV AD (OR=1.30, p=1.3×10^−13^) or DSM-5 AUD (OR=1.29, p=6.7×10^−13^) diagnoses. The association of PRS_SMK_ with FTND score was more significant (β=0.56, p=7.3×10^−49^) than with DSM-IV TD (OR=1.67, p=1.6×10^−46^). In contrast, the association of PRS_OUD_ with the DSM-5 OUD criterion count was marginally less significant (β=0.49, p=3.3×10^−15^) than the association with the DSM-IV OD (OR=1.28, p=2.9×10^−16^) and DSM-5 OUD (OR=1.28, p=3.1×10^−16^) diagnoses. PRS_CAN_ was only nominally associated with the respective criterion count (β=0.16, p=2.2×10^−4^) and DSM diagnoses (DSM-IV CD: OR=1.13, p=5.0×10^−4^; DSM-5 CUD: OR=1.12, p=3.0×10^−4^; Figure 2A, Supplementary Table 12).

### Associations with Phenotypes Involving the Same Substance

PRS that were associated with their respective substance use diagnosis were also associated with phenotypes for the same substance. PRS_AUD_ was associated with 8 alcohol phenotypes in AFR individuals (Figure 2B; Supplementary Table 6), and 32 alcohol phenotypes in EUR individuals (Figure 2B; Supplementary Table 9). In AFR, PRS_AUD_ was associated with 3 of the 4 criteria for alcohol abuse, including “continued use despite social/interpersonal problems” (OR=1.20, p=1.1×10^−7^), which was more significantly associated with PRS_AUD_ than the diagnosis itself. PRS_AUD_ was also significantly associated with frequent alcohol use, alcohol abuse, “ever had blackout”, and 2 of the 7 DSM-IV AD criteria (“unsuccessful efforts to decrease use” and “used more than intended”). In EUR, PRS_AUD_ was associated with all individual AUD diagnostic criteria and with “sought treatment”, frequent use, age of first use, “ever had blackout”, 9 withdrawal symptoms (e.g., “depressed mood”) and 6 symptoms related to heavy use (e.g., “depression”).

Among EUR individuals, PRS_OUD_ was associated with 41 opioid phenotypes (Figure 2B; Supplementary Table 10), including 2 associations that were more significant than with the diagnosis: “time spent obtaining/using” (OR=1.30, p=5.34×10^−18^) and “ever used opioids” (OR=1.28, p=1.9×10^−16^). PRS_OUD_ was significantly associated with 10 OD and abuse criteria, all except for “legal problems,” with which there was only a nominal association (OR=1.10, p=0.02). PRS_OUD_ was also significantly associated with “sought treatment”, frequent use, 4 symptoms of heavy use, and 18 withdrawal symptoms, the most significant of which was “depressed mood” (OR=1.26, p=3.7×10^−14^).

Among EUR individuals, PRS_SMK_ was associated with 22 tobacco phenotypes (Figure 2B; Supplementary Table 11), including all 7 TD criteria. Following the top associations with FTND score and the DSM-IV diagnosis of TD, the most significant association was “smoked over 100 cigarettes in lifetime” (OR=1.62, p=8.6×10^−45^). Associations were also found with frequent tobacco use, “sought use”, “ever used tobacco”, age at first use, symptom of use “health problems”, and 8 withdrawal symptoms, among which the greatest association was with “irritability” (OR=1.44, p=5.7×10^−33^).

Although only nominally associated with the diagnosis of CUD in EUR individuals, PRS_CAN_ (based on a lifetime measure of cannabis use) was significantly associated with 3 cannabis phenotypes (Figure 2B; Supplementary Table 12): “number of times used” (β=3.00, p=3.5×10^−7^), “ever used” (OR=1.20, p=1.3×10^−6^), and “regularly use” (OR=1.15, p=4.9×10^−6^).

### Phenome-wide Analyses

The PheWAS of PRS in AFR individuals identified no significant associations that passed Bonferroni correction in other phenotypic domains (Figure 3, Supplementary Tables 6-8). However, we identified multiple significant associations across phenotypic domains in EUR individuals (Figure 3, Supplementary Tables 9-12). For all 4 PRS in EUR individuals, the largest number of associations were with other substance use phenotypes. PRS_AUD_ was associated with 126 phenotypes in 12 categories, including phenotypes in all 7 substance use categories, the most significant of which was DSM-IV TD (OR=1.35, p=9.1×10^−19^) and “ever used cocaine” (OR=1.34, p=3.1×10^−18^). PRS_OUD_ was associated with 79 phenotypes in 12 categories, including those in all 7 substance use categories, the most significant of which were DSM-IV TD (OR=1.28, p=1.1×10^−14^), “sought treatment for cocaine use” (OR=1.25, p=7.7×10^−14^), and FTND score (β=0.27, p=1.4×10^−13^). PRS_SMK_ was associated with 160 phenotypes in 15 categories, including all 7 substance use categories. The most significant substance use phenotype was “ever used cocaine” (OR=1.47, p=5.1×10^−29^). PRS_CAN_ was associated with 24 phenotypes in 6 categories, though unlike the other PRSs, it was associated with phenotypes in only 4 of the 7 substance use categories. The most significant substance use phenotypes were “ever injected stimulants” (OR=1.19, p=6.1×10^−8^) and “ever used” stimulants (OR=1.18, p=6.6×10^−8^), hallucinogens (OR=1.18, p=8.8×10^−8^) or sedatives (OR=1.16, p=6.5×10^−7^).

**Figure 3.**
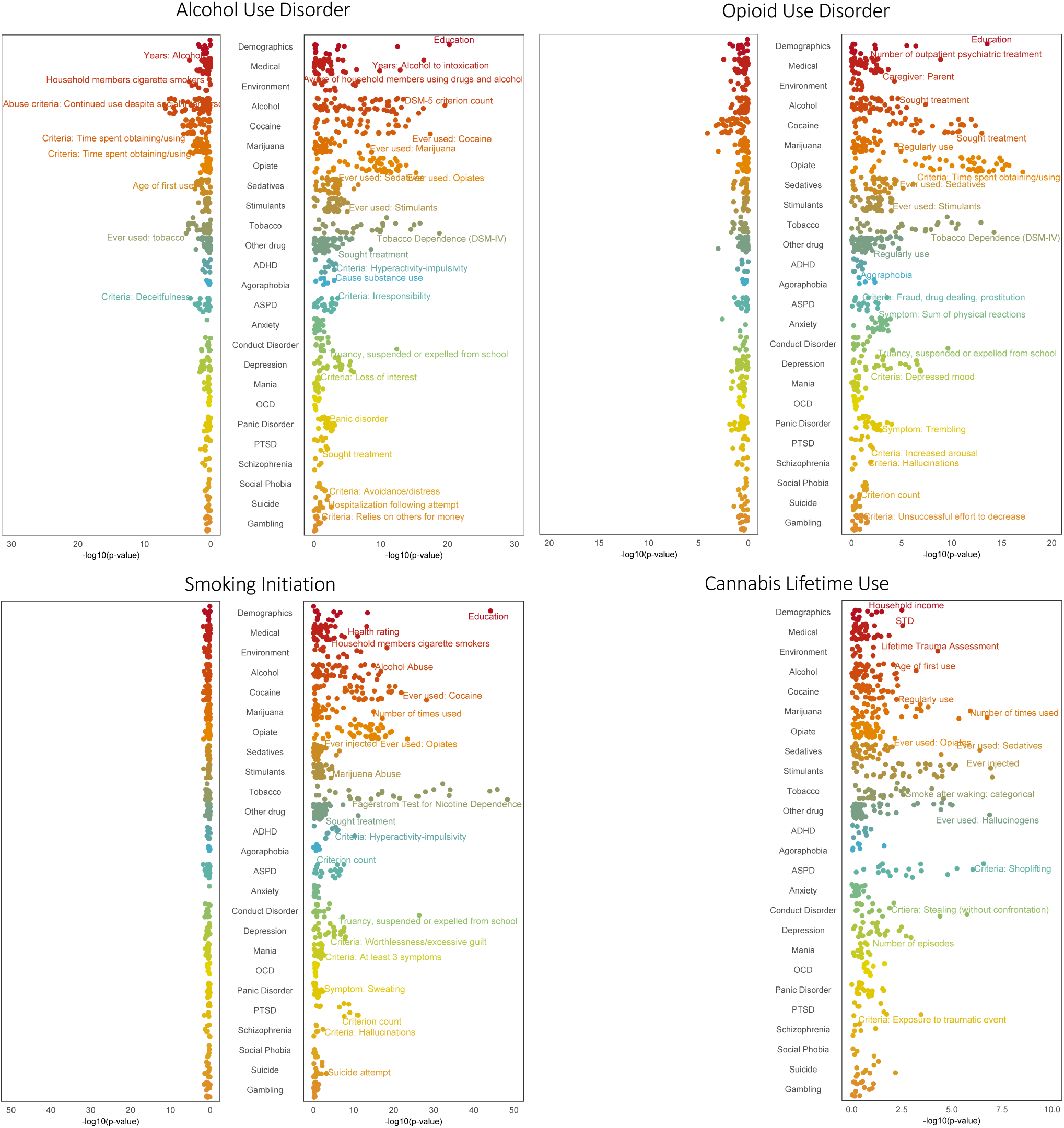
PheWAS of SUD PRS. PheWAS results for each PRS in AFR individuals (left panel of each graph) and EUR individuals (right panel of each graph). Within each category, the top associated phenotype was labelled if it passed FDR correction.

The psychiatric phenotype most significantly associated with PRS_AUD_, PRS_OUD_, and PRS_SMK_ was “truancy, suspended or expelled from school” in the conduct disorder domain (PRS_AUD_: OR=1.27, p=7.7×10^−13;^ PRS_OUD_: OR=1.22, p=4.5×10^−10^; PRS_SMK_: OR=1.44, p=7.9×10^−27^). PRS_AUD_ and PRS_OUD_ were both associated with multiple depression-related phenotypes, including the MDD criterion count (PRS_AUD_: β=0.24, p=4.5×10^−6^; PRS_OUD_: β=0.25, p=5.3×10^−7^) and MDD diagnosis (PRS_AUD_: OR=1.14, p=8.7×10^−6^; PRS_OUD_: OR=1.16, p=4.2×10^−7^). The second most significant phenotype for PRS_SMK_ was the criterion count for PTSD (β=0.18, p=4.0×10^−12^). PRS_SMK_ was also associated with phenotypes for depression, ASPD, and ADHD. PRS_CAN_ was associated with 4 phenotypes in the ASPD domain, including the ASPD diagnosis (OR=1.20, p=2.0×10^−5^), and 2 in the conduct disorder domain: “stealing (without confrontation)” (OR=1.18, p=3.4×10^−6^) and “persistent pattern of behavior” (OR=1.18, p=2.3×10^−6^).

For PRS_AUD_, PRS_OUD_, and PRS_SMK_, the most significant association with a demographic phenotype was a negative association with educational attainment (PRS_AUD_: β=-0.21, p=3.2×10^−21^; PRS_OUD_: β=-0.16, p=1.7×10^−14^; PRS_SMK_: β=-0.31, p=3.8×10^−45^). These 3 PRS were also negatively associated with household income and positively associated with the number of outpatient psychiatric treatments. PRS_AUD_ and PRS_SMK_ were both positively associated with childhood environmental variables, including “aware of household members using drugs or alcohol” and “frequent use of drugs/alcohol in household”. PRS_AUD_ was also associated with relevant phenotypes in the medical section, such as “alcohol used to intoxication” (β=0.95, p=2.5×10^−17^) and “health rating” (higher score = poorer health; β=0.08, p=2.4×10^−7^). PRS_SMK_ and PRS_CAN_ were positively associated with the lifetime trauma assessment (PRS_SMK_: OR=1.18, p=3.3×10^−8^; PRS_CAN_: OR=1.13, p=3.4×10^−5^) and PRS_OUD_ was negatively associated with having had a parent as the main caregiver (PRS_CAN_: OR=0.84, p=5.5×10^−5^).

## Discussion

PheWAS is a valuable tool for exploring cross-trait associations of phenotypes with genetic liability for specific disorders. However, most PheWAS conducted to date have used EHR data, which are typically limited to high-level phenotypes based on clinical diagnosis. Here, we describe the development of a dataset for PheWAS derived from the Yale-Penn sample, ascertained using a detailed psychiatric interview whose administration included multiple quality control procedures(13). We selected features to reduce the number of variables from 3,727 to the 685 that we considered informative for genetic analysis. We further refined cases and controls for each binary variable by applying similar methodology to that commonly used in EHR PheWAS(9), removing subthreshold individuals who met ≧1 criteria, but not the full diagnosis. Our approach yielded novel phenotypic associations with PRS for 4 substance use traits, particularly for subthreshold criteria or pertinent symptoms available only in a deeply-phenotyped sample such as that derived from the Yale-Penn study.

The SSADDA has multiple advantages as an assessment tool. Diagnoses made using a semi-structured interview following careful training procedures, with pre-specified criteria and strict quality control methods, have been shown to yield valid diagnoses(30) that likely are more accurate than those derived from EHR billing codes. As expected, many of the PRSs were associated with their respective primary diagnoses, providing a measure of validation of the approach. Likely due to the comparatively small GWAS discovery samples, PRS_CAN_ was not associated with the primary diagnoses; and, despite similar target sample sizes in the AFR and EUR individuals, few associations were identified for all PRS in the AFR sample. This reflects the lack of power for AFR in the parent GWAS studies, due to the much smaller AFR sample size.

The detailed information obtained with the SSADDA makes it possible to evaluate the impact of substance use genetic risk on a variety of substance-related traits not typically available in EHRs. For example, we found associations of tolerance and withdrawal – the 2 physiological SUD criteria. A genetic predisposition for tolerance could be informative for prevention efforts and a genetic propensity to experience withdrawal symptoms could help to identify individuals at high risk for severe symptoms to permit early intervention to limit symptom progression. In AFR, the criterion most strongly associated with PRS_AUD_ was “continued use despite social/interpersonal problems”, which was the second strongest association in EUR. Although in DSM-IV, this is an alcohol abuse criterion, factor analysis has shown that it loads on the same factor as the 7 alcohol dependence criteria and in item response theory analysis it is among those with the greatest information value(31). The strength of association with this criterion reflects impaired control, a central element in the alcohol dependence syndrome construct first described by Edwards and Gross(32). The DSM-IV substance abuse criterion “legal problems” had the fewest significant associations with any of the respective PRS. This is consistent with the results of twin and epidemiologic studies in which the legal criterion has the lowest loading of the DSM-IV criteria and low discriminatory power(33,34), supporting its omission from DSM-5(35). Our results also showed several associations with craving, a criterion that was added in the transition from DSM-IV to DSM-5. Although craving is not reported by all individuals with SUDs, in some studies it is predictive of relapse(36,37) and has been a target of pharmacological and psychosocial treatment(38–40).

In addition to the association of PRS with the phenotypes affiliated with the primary substance, each PRS also showed multiple associations with other substance use traits. The high genetic correlation among substance use traits, demonstrated by the discovery GWAS for each of these traits(2–6), suggests that this likely reflects true shared genetic effects. However, given the high levels of substance-related comorbidity in the Yale-Penn sample, the results likely also reflect phenotypic correlation and ascertainment bias.

We replicated associations between genetic risk for SUDs and other traits, including psychiatric diagnoses. The association between AUD and major depressive disorder has been identified in PheWAS of AUD PRS(10) and problematic alcohol use PRS(6) and in the analysis of genetic correlations between AUD and depression(6). Here we dissect this further by identifying associations between PRS_AUD_ and PRS_OUD_ with specific features of major depression, including low mood and difficulty concentrating. The psychiatric-related phenotype most significantly associated with PRS_AUD_, PRS_OUD_, and PRS_SMK_ was “truancy, suspended or expelled from school” in the conduct disorder domain. Analysis of a longitudinal sample of youth showed that truancy significantly predicted the initiation of alcohol, tobacco, and cannabis use(41). Its association with substance-related PRS suggests a common factor underlying both truancy and initiation of different substances. In alignment with previous phenotypic studies showing positive correlations between substance use and antisocial behaviors(42), we also observed significant associations for all 4 EUR PRS with other criteria of antisocial personality disorder and conduct disorder, such as shoplifting, fraud, and cheating. Previous studies have identified genetic correlations between PTSD and SUDs(43). Despite the low prevalence of PTSD in this sample, among EUR subjects we observed a significant association of PRS_SMK_ with PTSD criterion count, 5 of the 6 individual PTSD criteria, lifetime trauma assessment, and seeking treatment for PTSD. Similarly, PRS_CAN_ was significantly associated with lifetime trauma assessment. These relationships help to elucidate the features that underlie these common co-occurring symptoms and disorders.

The SSADDA also assesses pertinent environmental variables. In EUR individuals, PRS_AUD_ and PRS_SMK_ were both positively associated with childhood environmental variables, including “aware of household members using drugs or alcohol” and “frequent use of drugs/alcohol in household,” whereas PRS_OUD_ was negatively associated with having a parent as the main caregiver. Such relationships are expected as these variables capture aspects of family history of substance use. However, although a family history of a SUD is associated with many substance use outcomes, it is not wholly overlapping with genetic risk and it has been argued that these 2 sources of information should be used together to provide a fuller measure of risk(44). These findings raise an important theoretical question that cannot be answered with the data available here: namely, do associations of PRS with features such as trauma, truancy, educational, and parental substance reflect intergenerational effects (i.e., “genetic nurture”?) or do they refute typical assumptions that the genetic and environmental components in GxE interactions are uncorrelated (i.e., adverse environments are evenly distributed across the range of PRS)?

Although the data from the Yale-Penn sample are more granular than those available from EHR-derived samples, the sample is comparatively small, as the recruitment and ascertainment activities are time consuming and resource intensive and thus less amenable to gene discovery than “big-data samples.” Thus, we believe that the strategy described here is complementary to the use of biobank data. However, interview-based studies such as this depend on individuals’ self-report, which is subject to recall bias and underreporting (although our recruitment strategy, which depended on self-report of an SUD, worked against this bias). Unlike EHR-based genetic studies, our study is cross-sectional and therefore lacks a longitudinal perspective. We were able to calculate PRS and conduct PheWAS in both AFR and EUR ancestry individuals by selecting the majority of the GWAS from the Million Veteran Program (MVP), a large and diverse biobank. However, MVP is comprised of veterans who are predominantly male and older and who have high rates of medical comorbidity, and thus differ from the target sample.

GWAS have begun to elucidate the high degree of genetic overlap among SUDs and related traits, though they are limited by a lack of deep phenotyping, a tradeoff with the large samples needed to provide adequate statistical power to identify common variants of small effect. This study relied on a carefully constructed diagnostic interview that enabled us to conduct analyses of primary substance use traits, results of which validated the effort, and a wealth of phenotypic data not captured in EHR-based biobanks (e.g., individual diagnostic criteria and symptoms, age of onset, and environmental variables). The continued growth of biobanks of increasing ancestral diversity will provide opportunities to compare the performance of PRS with the findings reported here. Additional enriched, deeply phenotyped samples will be needed to support such efforts.

## Data Availability

All data produced in the present work are contained in the manuscript.

## Acknowledgments

This study was supported by NIH grants AA028292 (to RLK), DA046345, and AA026364 and the Veterans Integrated Service Network 4 Mental Illness Research, Education and Clinical Center.

## Disclosures

Dr. Kranzler is a member of advisory boards for Dicerna Pharmaceuticals, Sophrosyne Pharmaceuticals, and Enthion Pharmaceuticals; a consultant to Sobrera Pharmaceuticals; the recipient of research funding and medication supplies for an investigator-initiated study from Alkermes; a member of the American Society of Clinical Psychopharmacology’s Alcohol Clinical Trials Initiative, which was supported in the last three years by Alkermes, Dicerna, Ethypharm, Lundbeck, Mitsubishi, and Otsuka. Drs. Kranzler and Gelernter are named as inventors on PCT patent application #15/878,640 entitled: “Genotype-guided dosing of opioid agonists,” filed January 24, 2018. The other authors have no disclosures to make.

## Notes

### Author Declarations

Yale-Penn participants were recruited at 5 sites (Yale University School of Medicine (APT Foundation; New Haven, CT), the University of Connecticut Health Center (Farmington, CT), the University of Pennsylvania Perelman School of Medicine (Philadelphia, PA), the Medical University of South Carolina (Charleston, SC), and McLean Hospital (Belmont, MA) in the United States for genetic studies of cocaine, opioid, and alcohol dependence. The study was approved by the institutional review board at each site and all participants gave written informed consent prior to data collection.

