## Supplementary Methods for "Phenome-wide association analysis of substance use disorders in a deeply phenotyped sample"

Variable selection

We retained demographic variables, including age, self-reported race and ethnicity, marital status, annual income level, educational attainment, and number of children. The retained medical variables included a history of a variety of physical conditions (e.g., cancer, asthma, migraine, heart conditions), hospitalizations and the reasons for them, history of medication use (e.g., antidepressants, antipsychotics, diabetes medications), psychiatric treatment history, and a self-rating of overall health.

The medical section is followed by sections that cover the use of alcohol, cocaine, opioids, and tobacco and a general substance use section that includes similar questions on cannabinoids, stimulants, sedatives, and other drugs. Across all substance use sections, we retained measures of whether the respondent ever used the substance and, if so, the age of first use, measures of quantity and frequency of use, route of administration, DSM-IV and DSM-5 criteria (DSM-5 diagnosis was possible due to the inclusion of an item assessing craving), other pertinent symptoms (e.g., physiological and psychological withdrawal symptoms), treatment history, and age of onset of a disorder. For all SUDs, we generated diagnoses based on DSM-IV (i.e., abuse and dependence) and DSM-5 (i.e., SUD: mild, moderate, or severe) criteria and criterion counts for both diagnostic systems, using a summary score for all 11 criteria in each system. The exception to this was tobacco, where only the DSM-IV diagnosis could be generated; we therefore also retained the responses to the 6 items of the Fagerström Test for Nicotine Dependence (FTND)^1^.

We used a similar approach to create consistent measures across psychiatric disorders, retaining symptoms that underpin the DSM criteria, other pertinent symptoms, DSM-IV diagnoses, criterion counts, age of onset of symptoms and diagnosis, treatment received, and length and number of episodes. The SSADDA interview also contains a section on suicidality. We retained items that assessed the age of onset and characteristics of suicidal ideation, suicide plans, and suicide attempts. For individuals who endorsed having made a suicide attempt, we retained information on the number of attempts, age of first attempt, treatment received, and the severity of suicidal intent. From the environment section, we retained information on childhood experiences and events, who the individual’s primary caregiver was, parental deaths, the number of household moves, exposures to violence and physical and sexual abuse, and whether the respondent observed adults using substances. Lifetime trauma exposure and childhood adversity variables were calculated using methods defined previously^2,3^.

Data cleaning

We used Python to extract and clean the phenotype data. First, the data were recoded as necessary to ensure consistency across all variables. For responses to binary questions, we coded all values to yield 3 possible response categories: “Yes”, “No” and “No Answer”. Each diagnostic section begins with a series of skip out questions. Individuals whose answers to these questions yield no positive diagnostic criteria skip out of the rest of the section. If a participant skips out of the section due to answering “No” to the skip out questions, we considered them an unexposed control for the purposes of binary phenotypes within that section. Continuous variables coded as “unsure” were removed. To eliminate the effects of extreme values in the continuous variables, we applied winsorization. Variables were winsorized with cut-off points at mean +/- 2*SD or mean +/- SD depending on the variable's distribution.

Genotyping and Imputation

Individuals were removed if genotype call rate <98%, and single nucleotide polymorphisms (SNPs) were removed if the missing rate >2%, minor allele frequency (MAF) <1%, or Hardy-Weinberg equilibrium p<1x10^-6^. Genotype data were imputed using the Michigan Imputation Server^4^ with the 1000 Genomes Phase 3 reference panel^5^. Imputed genotype data were generated for 13,487 individuals.

Following imputation, we extracted SNPs present in all 3 batches (N=8,820,767) and merged the datasets. We removed individuals with genotype call rate <95%, and retained SNPs with INFO scores >0.7, missing genotypes <1%, and MAF ≥1%. We inferred genetic ancestry for all individuals using a process described previously^6^. In brief, we used SNPs common to both the Yale-Penn dataset and the 1000 Genomes phase 3 reference panels to calculate principal components using PLINK 1.9^7^. The population closest to the subject based on the distance of 10 PCs was assigned to the subject (AFR: N=6,286; EUR: N=6,460; Other=739). To remedy batch effects observed in PCs between the Yale-Penn III data and the Yale-Penn I and Yale-Penn II data, we removed SNPs with an allele frequency difference >0.04 among the 3 batches. We randomly removed 1 member of each pair of related individuals (pi_hat > 0.25) randomly from the data, leaving 4,918 AFR subjects and 5,692 EUR subjects for analyses (Supplementary Figure 1).
