## Supplementary figures and images for "Phenome-wide association analysis of substance use disorders in a deeply phenotyped sample"

### Supplementary Figures

Supplementary Figure 1

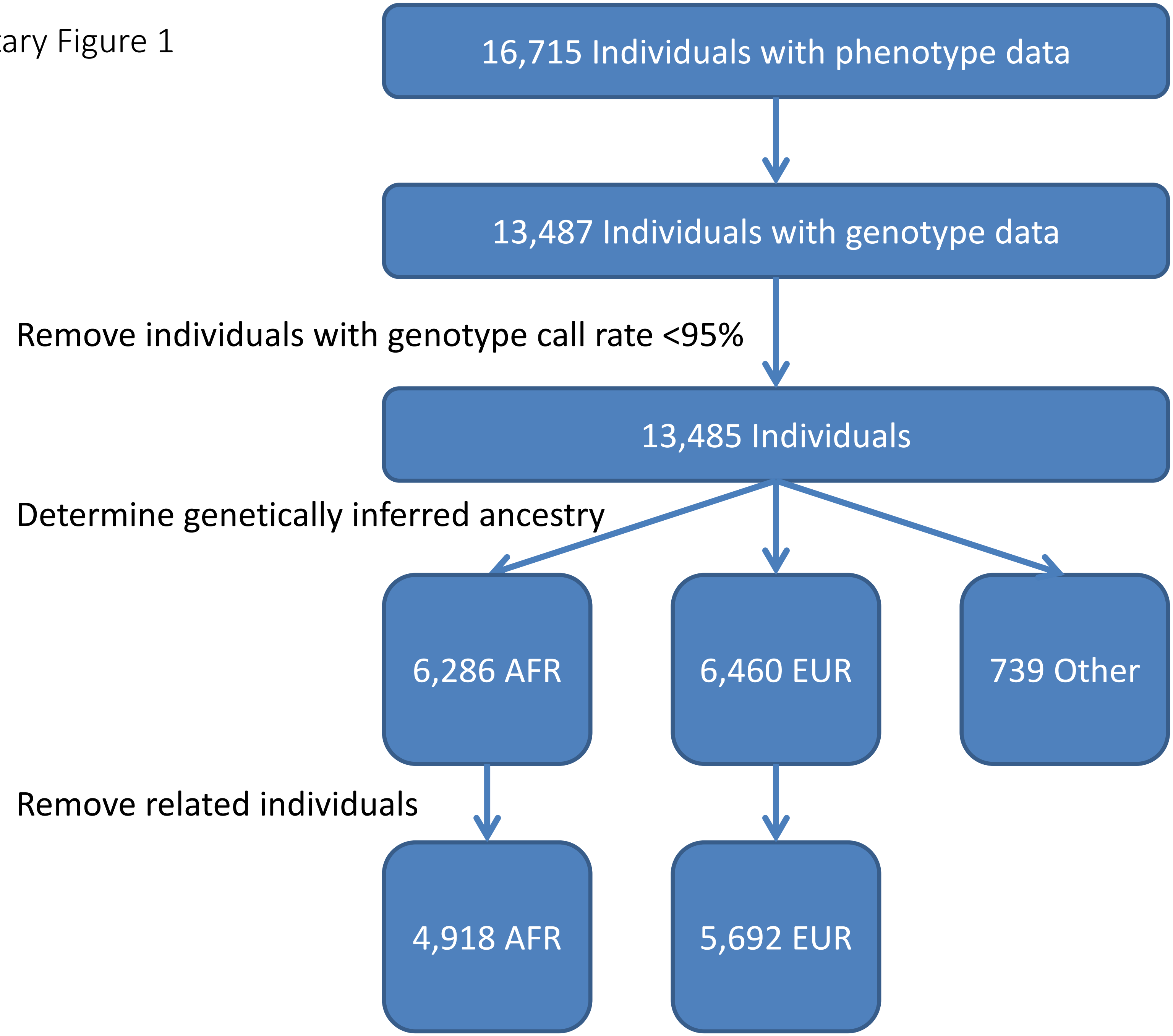
